# The Distribution of Optimism Across Sociodemographic Groups in 22 Countries

**DOI:** 10.1101/2025.03.14.25323964

**Authors:** Ying Chen, Laura D. Kubzansky, Eric S. Kim, Hayami Koga, Koichiro Shiba, R. Noah Padgett, Renae Wilkinson, Byron R. Johnson, Tyler J. VanderWeele

## Abstract

Prior research (mainly from Western industrialized countries) documents associations between greater dispositional optimism (a generalized expectation that good things will happen) and improved health and well-being. However, less is known about whether and how levels of optimism differ across countries and across sociodemographic groups within different countries. This study presents a cross-national exploration of optimism, and its variations across sociodemographic groups. Using a sample of 202,898 adults from 22 diverse countries, we examined the relationships between optimism and key sociodemographic factors in each country separately, and cross-nationally by pooling results across countries using meta-analytic techniques. Our results suggest that mean optimism levels vary substantially across countries. Optimism also varies significantly across most of the sociodemographic factors included in our analyses. In the pooled results across countries, individuals who are older, female, married, employed, highly educated, attending religious services frequently, and native-born reported higher mean optimism levels. In the country-specific analyses, the sociodemographic variation in optimism differs across countries, indicating diverse societal influences. The findings of this study provide novel insights into the population distribution of optimism and disparities in optimism by sociodemographic groups across countries. This study provides a valuable foundation for future investigations into sociocultural influences that shape optimism.

## INTRODUCTION

Evidence on the importance of well-being has expanded drastically over recent years^1^. Governments worldwide have been increasingly interested in identifying targets for strategies that aim at enhancing population well-being^2^. A facet of psychological functioning that has been linked to multiple aspects of well-being is dispositional optimism, which is often defined as “a generalized expectation that good things will happen”^3^. While there are various forms of optimism such as “resourced optimism” (optimism that is grounded in one’s resources [e.g., education, financial resources, etc.]) and “agentive optimism” (optimism that is grounded in one’s commitment and personal agency in pursuing the desired outcomes), most forms of optimism involve a positive expectation about the future at their core and can be classed into a genus called “expectancy optimism”^4^. The present study focuses on this generic form of expectancy optimism.

According to one of the major theoretical frameworks for optimism, the Expectancy-Value Theory, individuals with a higher level of optimism are more likely to feel confident in attaining desired outcomes and persist in goal-directed efforts even in difficult situations, which leads to greater well-being and resilience^3^. A body of empirical literature, mostly based on data from United States and European countries supports this premise, reliably finding that optimism is associated with better health and well-being among adults. For instance, greater optimism has been linked with lower risks of mortality^5^, cardiovascular diseases^6^, and mental illness^7^, and with healthier lifestyles^8^. Optimism has less often been studied in non-Western cultures, but preliminary cross-sectional evidence indicates that the correlations between one’s expectation about future life and well-being may vary across countries^9^. Such variations might be attributable to differential effects of optimism on health in part due to differences in social contexts. However, compared to the health effects of optimism, the social distribution of optimism is less well-understood. A comprehensive understanding of the sociodemographic distribution of optimism across societies is essential for identifying social groups for whom interventions to enhance optimism may be most meaningful.

The population mean levels of optimism may vary by country. For instance, differences in cultural values and social norms between countries may shape the extent to which people adopt and maintain an optimistic mindset at the population level^9^. Specifically, individualistic cultures value attaining personal goals, which may motivate people to persist in goal-directed behaviors and take an optimistic outlook for future outcomes. In comparison, collectivist cultures prioritize harmonious interpersonal relationships over self-enhancement, which may lead to less emphasis on cultivating one’s own expectations and developing an optimistic outlook^10,11^. With regard to empirical work, some earlier studies found that people in Asian countries are less optimistic and have less of an inclination toward positive thinking than those in North America and Europe^12,13^. However, recent studies using large-scale cross-national data (including a study of 150,048 participants from 142 countries^9^, a study of 15,185 people from 61 countries^14^, and a meta-analysis of 89,000 participants from 22 countries^15^) suggested that people in most countries are generally more optimistic than not. These prior studies are, nevertheless, subject to several methodological limitations, such as the non-representative samples and the use of measures for other psychological factors as proxies for optimism, which limit the validity of the findings. Overall, the extent to which mean levels of optimism vary by country remains unclear.

Within each country, optimism may not be equally distributed across sociodemographic groups that often have differential access to resources. Disparities-related structural and social factors may provide a context for fostering or restricting an optimistic mindset. First, optimism may be patterned by age groups. Prior evidence suggests that older adults on average have a more optimistic outlook than younger adults in some Western countries (e.g., United States, Canada), whereas optimism decreases with age or is unrelated to age in non-Western cultures (e.g., China)^13^. Second, empirical findings on gender and optimism are mixed. Some studies found that men are more optimistic than women in the context of future economic situations^16^, whereas other studies found little gender difference in general optimism^17^. There is also growing evidence from Western countries suggesting that gender minorities have higher risks of mental distress (e.g., depression, anxiety, suicidality)^18^, but to our knowledge optimism among gender minorities has seldom been studied. Third, prior work suggests that people’s level of optimism may vary by marital status. For instance, those who are married or have a partner on average reported a higher level of optimism compared to those who are widowed^19,20^. Fourth, individuals’ socioeconomic status (SES) may shape their outlook for the future. For instance, a study in American adults found that individuals with higher (vs. lower) SES consistently reported higher optimism across multiple indicators of SES, including educational attainment, occupational prestige, and household income^17^. Beyond SES disparities, some social identity factors (e.g., race/ethnicity, immigration status) may also be associated with psychosocial stress (e.g., discrimination, marginalization) that shapes one’s outlook to the future^21^. Fifth, a sense optimism is highly valued in many religious communities around the world^22^. Data from the United States suggests that greater religious participation is on average associated with higher levels of optimism^23,24^, although the dynamics may vary across contexts^25^.

Prior studies have significantly advanced our understanding of optimism across countries and its sociodemographic variation, but some knowledge gaps remain. First, many studies used non-representative samples, which limits the external validity of study findings even within the countries in which these studies were conducted. Second, prior studies seldom examined population distribution of optimism through a lens of disparities. Specifically, most studies have focused on population averages of optimism, whereas inequalities in its distribution within societies are less understood. Evaluating social disparities in optimism will help inform a more equitable promotion of optimism and population well-being. Third, most studies were conducted using samples from North American and European countries, whereas the population mean levels and the social patterning of optimism in other parts of the world is less understood. Given cultural values and societal-level factors (e.g., economic development, income inequality, institutional trust) are likely to shape the extent to which individuals will cultivate an optimistic mindset^13,14^, we cannot assume that distributions evident in North America and Europe will be similar across the world.

To begin addressing these knowledge gaps, using data from 202,898 adults in 22 countries, the present study examined 1) mean levels of optimism and overall inequalities in the population distribution of optimism (evaluated by Gini coefficient, see details in the Methods section) in each country, 2) mean levels of optimism by major sociodemographic factors (including age, gender, marital status, education, employment status, immigration status, and religious service attendance; we also examined religious affiliation and race/ethnicity in countries wherever data were available) on average across countries, and 3) mean levels of optimism by sociodemographic factors within each country separately. We hypothesized that 1) mean levels of optimism would vary across countries and that optimism would not be equally distributed within each country, 2) mean levels of optimism would differ by sociodemographic groups on average across countries, and 3) the extent to which mean levels of optimism differ by sociodemographic groups would also vary by country.

## METHODS

This study used Wave 1 data from the Global Flourishing Study (GFS). The description of the methods below has been adapted from VanderWeele et al.^26^ (the paper by VanderWeele et al. is scheduled to be published before the present study, and the reference will be updated when it becomes available). Further methodological detail of the GFS is available elsewhere^27–32^. The present study is a secondary analysis of the existing GFS data.

### Study Population

The Global Flourishing Study is a longitudinal study that enrolled 202,898 adults (age range: 18 to 99 years) from 22 culturally and geographically diverse countries, with the samples weighted to be nationally representative within each country. Survey items queried multiple aspects of well-being such as happiness, health, having a sense of meaning and purpose, character, social relationships, and financial stability, along with a broad range of demographic, socioeconomic, political, religious, personality, childhood, community, and behavior factors.

Cognitive interview and pilot-testing were conducted to validate the translation of survey questionnaires, and the translations were then sent to scholars in participating countries for further evaluation^28^. Data collection was carried out by Gallup via a combination of modes (e.g., in-person, phone, web) that varied across countries^31^. Data for Wave 1 were collected principally during 2023, while some countries began data collection in 2022^27^. The following countries/territories were included in Wave 1 data collection: Argentina, Australia, Brazil, Egypt, Germany, Hong Kong (Special Administrative Region of China), India, Indonesia, Israel, Japan, Kenya, Mexico, Nigeria, the Philippines, Poland, South Africa, Spain, Sweden, Tanzania, Turkey, United Kingdom, and the United States. These countries were chosen to 1) maximize coverage of the world’s population, 2) ensure geographic, cultural, and religious diversity, and 3) prioritize feasibility and existing data collection infrastructure. The precise sampling design to ensure nationally representative samples varied by country^27^. The Wave 1 response rates across countries ranged from 27% to 100% for probability-based samples and from 60.5% to 100% for non-probability-based samples^27^. The data are publicly available through the Center for Open Science (COS, https://www.cos.io/gfs). Details about the GFS study methodology and survey development were reported elsewhere^27,28^.

The present study used data from all participants in Wave 1 of GFS (N=202,898). Poststratification and nonresponse adjustments were performed to ensure the sample was representative of the adult population in each country^27,31^. Ethical approval was granted by the institutional review boards at Baylor University and Gallup, and all participants provided informed consent. All methods were carried out in accordance with relevant ethical guidelines and regulations.

### Assessment of Optimism

One item from the previously validated Revised Life Orientation Test^33^ was used to measure optimism: “Overall, I expect more good things to happen to me than bad”. The item was selected by GFS based on inputs from experts in optimism, and cognitive interviews and pilot testing were conducted by GFS to evaluate the performance of the item^28,29^. The response was rated on a Likert scale ranging from 0 (strongly disagree) to 10 (strongly agree). The response was analyzed as a continuous score, with a higher score indicating a higher level of optimism.

### Assessment of Sociodemographic Characteristics

Sociodemographic factors were assessed through participant self-reports. These included age (in years, responses grouped into “18-24’, “25-29”, “30-39”, “40-49”, “50-59”, “60-69”, “70-79”, and “80+”), gender (male, female, or other), marital status (single/never married, married, separated, divorced, widowed, or domestic partner), employment status (employed by an outside organization, self-employed, retired, student, homemaker, unemployed and searching, or other), educational attainment (up to 8 years, 9-15 years, or 16+ years, following the categorization approach for educational attainment in Gallup World Poll annual global survey^34^), and immigration status (born in this country, born in another country). Because a large proportion of the world population identified with a religious group and researchers have argued that religion is a social determinant of health^35,36^, we additionally included religious service attendance (>once/week, once/week, one-to-three times/month, a few times/year, or never) as a sociodemographic factor in this study. We also assessed religious affiliation, and response options included 15 major religions [e.g., Christianity, Islam, Hinduism, Buddhism, Judaism, etc.], “some other religion”, or “No religion/Atheist/Agnostic”; response categories varied by country^37^. Race/ethnicity was assessed in most countries (except in Germany, Japan, Spain, and Sweden, due to restrictions in collecting such data), with response options varied as appropriate for each country.

### Statistical Analyses

The descriptive analyses show participant demographic characteristics in individual countries and in the total sample, weighted to be nationally representative within each country. Mean levels of optimism (adjusted for complex survey design and weighted to be nationally representative) were estimated for each country separately along with 95% confidence intervals, standard deviations, and for presentation purposes, we ordered the mean estimates from highest to lowest. Additionally, we calculated Gini coefficient^38^ to evaluate overall inequality in the population distribution of optimism within each country. Gini coefficient, ranging from 0 (perfect equality) to 1(highest inequality), was originally used to assess population income inequality and has been increasingly used in measuring disparities in health and well-being^39^. Variation in mean optimism by sociodemographic categories was also estimated, with all analyses initially conducted within country (reported in online supplement). A global (joint) test was conducted (with a global p-value calculated) to assess whether optimism levels varied statistically significantly across categories of each sociodemographic variable within each country.

In the primary analyses we conducted random effects meta-analysis to pool mean values of optimism in each specific sociodemographic category from the country-specific analyses^40,41^, along with 95% confidence intervals, standard errors, lower and upper limits of 95% prediction intervals, heterogeneity (τ), and I^2^ to estimate the extent to which optimism levels in a given sociodemographic category vary across countries^42^. Forest plots of estimates are shown in the online supplement. We examined religious affiliation and race/ethnicity (when available) in country-specific analyses but not with the meta-analyses, because the observed response categories for these two demographic factors varied by country. We reported a pooled global p-value^43^ across countries to evaluate if optimism levels varied statistically significantly by each sociodemographic variable within at least one country. Bonferroni corrected p-value thresholds are provided in the meta-analytic results based on the number of sociodemographic variables^44,45^.

Meta-analyses were conducted in R (R Core Team, 2024) using the metafor package^46^, and country-specific analyses were performed using SAS 9.4 (SAS Institute, Inc). All statistical tests were 2-sided. As a supplementary analysis, population weighted meta-analysis was performed to pool results from country-specific analyses. All analyses were pre-registered with COS prior to data access (https://doi.org/10.17605/OSF.IO/WDSX2); all code to reproduce analyses are openly available in an online repository^32^.

### Missing Data

In the total sample combined across countries, 0.41% of participants had missing data on optimism, and missing data on sociodemographic factors ranged from 0.01% to 1.01% (except race/ethnicity, which was not measured in some countries). We conducted multivariate imputation by chained equations (5 imputed datasets created) to impute missing data on all variables in each country separately^47–49^. We included sampling weights in the imputation model to account for missingness related to probability of inclusion^50^. As a sensitivity analysis, we also reran the primary analyses using complete-case analyses (without imputation for missing data).

### Accounting for Complex Sampling Design

The GFS used different sampling schemes across countries based on availability of existing panels and recruitment needs^27^. All analyses in this study accounted for the complex survey design components by including weights, primary sampling units, and strata. Additional methodological details are reported elsewhere^30,31^.

## RESULTS

### Descriptive Analyses

Table 1 shows participant characteristics in the weighted cross-national sample. There are relatively similar proportions of people in different age groups, except fewer participants were older than 70+ years (Table 1). The weighted sample has a balanced representation of female (51%) and male (49%) participants, and a small proportion who identified as other gender (0.3%). Higher proportions of participants were married (53%), employed (57%), had 9-15 years of education (57%) and were native-born (94%), with a smaller proportion reporting they never attended religious services (37%). Sample sizes in each country ranged from 1,473 (Turkey) to 38,312 (United States). Participant characteristics for each country are shown in Supplementary Table S1A to S22A.

**Table 1.** Distribution of the Sociodemographic Factors in the Overall Sample Combined Across 22 Countries (Weighted to be Nationally Representative within Each Country, N=202,898).

| Sociodemographic Factors | Category | N (%) |
| --- | --- | --- |
| Age group | 18-24 | 27,007 (13%) |
|  | 25-29 | 20,700 (10%) |
|  | 30-39 | 40,256 (20%) |
|  | 40-49 | 34,464 (17%) |
|  | 50-59 | 31,793 (16%) |
|  | 60-69 | 27,763 (14%) |
|  | 70-79 | 16,776 (8%) |
|  | 80+ | 4,119 (2%) |
|  | Missing | 20 (<1%) |
| Gender | Male | 98,411 (49%) |
|  | Female | 103,488 (51%) |
|  | Other | 602 (<1%) |
|  | Missing | 397 (<1%) |
| Marital status | Married | 107,354 (53%) |
|  | Separated | 5,195 (3%) |
|  | Divorced | 11,654 (6%) |
|  | Widowed | 9,823 (5%) |
|  | Domestic partner | 14,931 (7%) |
|  | Single/Never married | 52,115 (26%) |
|  | Missing | 1,826 (1%) |
| Employment status | Employed by an outside organization | 78,815 (39%) |
|  | Self-employed | 36,362 (18%) |
|  | Retired | 29,303 (14%) |
|  | Student | 10,726 (5%) |
|  | Homemaker | 21,677 (11%) |
|  | Unemployed and looking for a job | 16,790 (8%) |
|  | None of these/other | 8,431 (4%) |
|  | Missing | 793 (<1%) |
| Education | Up to 8 years | 45,078 (22%) |
|  | 9-15 years | 115,097 (57%) |
|  | 16+ years | 42,578 (21%) |
|  | Missing | 146 (<1%) |
| Religious service attendance | >1/week | 26,537 (13%) |
|  | 1/week | 39,157 (19%) |

| **Sociodemographic Factors** | **Category** | **N (%)** |
| --- | --- | --- |
|  | 1-3/month | 19,749 (10%) |
|  | A few times a year | 41,436 (20%) |
|  | Never | 75,297 (37%) |
|  | Missing | 722 (<1%) |
| Immigration status | Born in this country | 190,998 (94%) |
|  | Born in another country | 9,791 (5%) |
|  | Missing | 2,110 (1%) |
| Country | Argentina | 6,724 (3%) |
|  | Australia | 3,844 (2%) |
|  | Brazil | 13,204 (7%) |
|  | Egypt | 4,729 (2%) |
|  | Germany | 9,506 (5%) |
|  | Hong Kong (S.A.R of China) | 3,012 (1%) |
|  | India | 12,765 (6%) |
|  | Indonesia | 6,992 (3%) |
|  | Israel | 3,669 (2%) |
|  | Japan | 20,543 (10%) |
|  | Kenya | 11,389 (6%) |
|  | Mexico | 5,776 (3%) |
|  | Nigeria | 6,827 (3%) |
|  | Philippines | 5,292 (3%) |
|  | Poland | 10,389 (5%) |
|  | South Africa | 2,651 (1%) |
|  | Spain | 6,290 (3%) |
|  | Sweden | 15,068 (7%) |
|  | Tanzania | 9,075 (4%) |
|  | Turkey | 1,473 (1%) |
|  | United Kingdom | 5,368 (3%) |
|  | United States | 38,312 (19%) |

Note. S.A.R. = Special Administrative Region.

### Ordered Mean Level of Optimism by Country

Population mean scores of optimism are highest in Brazil (9.22 on a scale from 0 to 10, 95% confidence interval [CI]: 9.18, 9.25) and Indonesia (9.15, 95% CI: 9.10, 9.21), and lowest in Japan (6.74, 95% CI: 6.70, 6.77) and United Kingdom (6.99, 95% CI: 6.89, 7.08) (Table 2).

**Table 2.** Ordered Means of Optimism with Standard Deviations and Gini Coefficients.

| **Country** | **Mean** | **95% Confidence Interval** | **Standard Deviation** | **Gini\*** |
| --- | --- | --- | --- | --- |
| Brazil | 9.22 | (9.18,9.25) | 1.62 | 0.07 |
| Indonesia | 9.15 | (9.10,9.21) | 1.69 | 0.07 |
| Mexico | 9.06 | (9.00,9.12) | 1.66 | 0.08 |
| Kenya | 8.92 | (8.85,9.00) | 2.20 | 0.10 |
| Nigeria | 8.90 | (8.82,8.97) | 1.77 | 0.09 |
| Argentina | 8.86 | (8.79,8.92) | 1.83 | 0.09 |
| Philippines | 8.79 | (8.72,8.86) | 2.01 | 0.10 |
| Tanzania | 8.75 | (8.65,8.85) | 2.36 | 0.11 |
| Israel | 8.38 | (8.26,8.51) | 1.78 | 0.11 |
| Spain | 8.25 | (8.18,8.31) | 1.97 | 0.12 |
| South Africa | 8.22 | (8.09,8.34) | 2.22 | 0.14 |
| India | 8.14 | (8.07,8.22) | 2.90 | 0.17 |
| Poland | 7.72 | (7.61,7.83) | 1.83 | 0.13 |
| Egypt | 7.67 | (7.56,7.77) | 2.57 | 0.18 |
| Turkey | 7.66 | (7.50,7.81) | 2.49 | 0.17 |
| United States | 7.65 | (7.59,7.71) | 2.29 | 0.16 |
| Germany | 7.50 | (7.45,7.56) | 2.11 | 0.15 |
| Australia | 7.41 | (7.31,7.51) | 2.27 | 0.16 |
| Sweden | 7.30 | (7.25,7.34) | 2.32 | 0.17 |
| Hong Kong | 7.23 | (7.13,7.33) | 2.10 | 0.16 |
| United Kingdom | 6.99 | (6.89,7.08) | 2.44 | 0.19 |
| Japan | 6.74 | (6.70,6.77) | 2.27 | 0.19 |

\* In this study, Gini coefficient measures inequality in the population distribution of optimism, and ranges from 0 (perfect equality) to 1 (highest inequality).

Overall, the higher-ranking countries are prominently those that are considered more collective (e.g., Brazil, Indonesia, Philippine, Nigeria, Kenya; Japan is an exception, which has the lowest mean score of optimism), whereas many high-income and individualistic countries are featured prominently in the lower-ranking countries (e.g., United States, Germany, Sweden, United Kingdom). Variation within country in responses to the optimism item is highest in India (standard deviation [SD]=2.90) and lowest in Brazil (SD=1.62). Further, countries with a lower ranking on mean optimism also tend to have greater inequality in population distribution of optimism, as indicated by a larger Gini coefficient.

### Sociodemographic Variation in Optimism Levels: Pooled Estimates across Countries

The random effects meta-analysis that pooled results from country-specific analyses shows mean optimism is patterned by all sociodemographic factors in the total sample (Table 3, all global p-values are below the Bonferroni corrected significance level of p<.007). Specifically, older versus (vs.) younger participants reported incrementally higher levels of optimism on average across countries. For example, on average across countries, participants aged 80+ years reported a mean optimism score of 8.21 (95% CI: 7.87, 8.55) on a scale from 0-10, whereas participants aged between 18-24 years reported a mean optimism score of 7.97 (95% CI: 7.59, 8.35). Mean optimism among females (mean=8.19, 95% CI: 7.87, 8.51) is higher than in males (mean=8.04, 9% CI: 7.72, 8.36) by 0.15 points, and is substantially higher than among individuals who identified as other gender (mean=6.98, 95% CI: 6.01, 7.95) by 1.21 points.

**Table 3.**
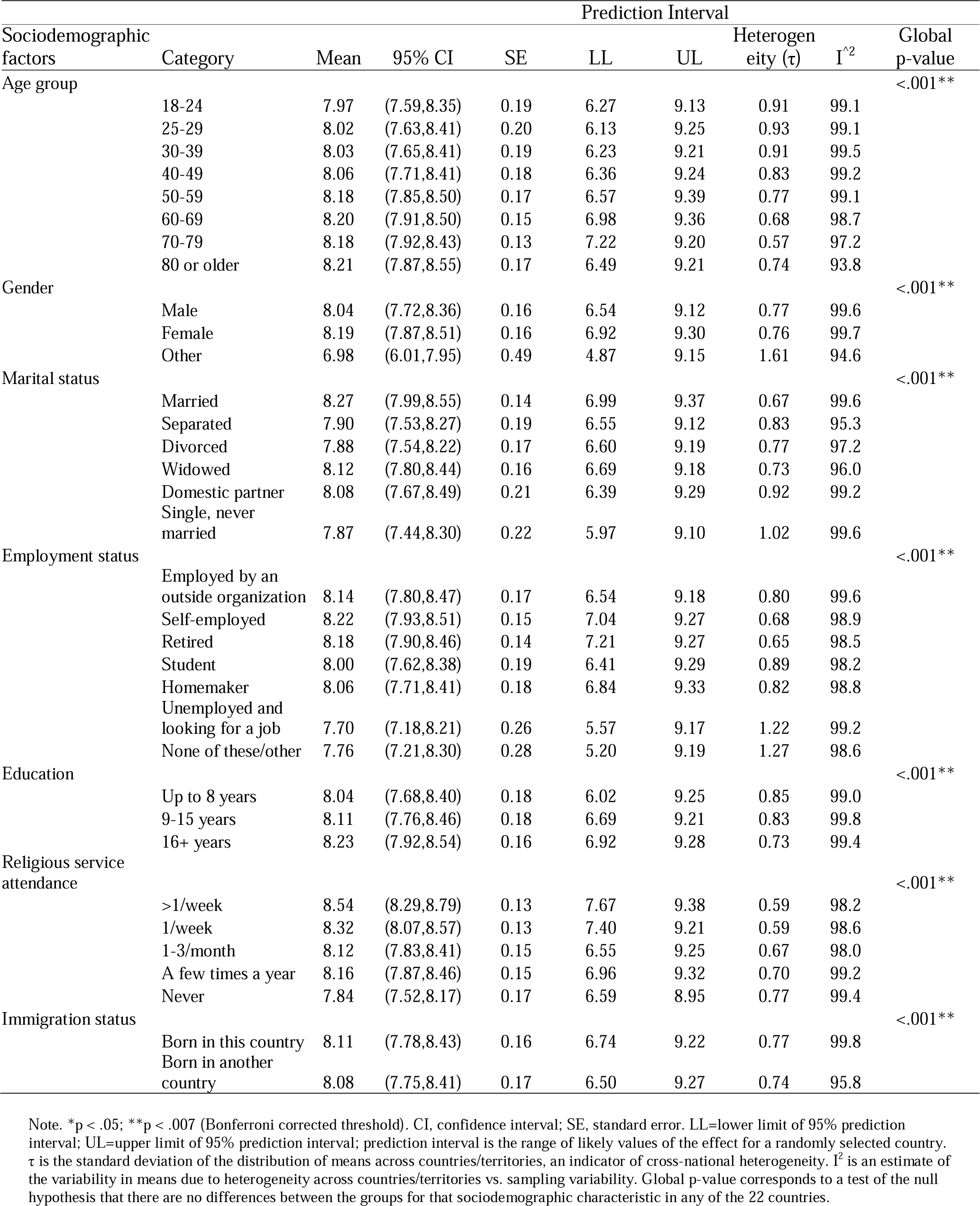
Random Effects Meta-Analysis of Mean Optimism Levels by Sociodemographic Category.

Further, those who are married (mean=8.27, 95% CI: 7.99, 8.55) reported a higher mean level of optimism than divorced (mean=7.88, 95% CI: 7.54, 8.22) or single/never married (mean=7.87, 95% CI: 7.44, 8.30) participants by approximately 0.40 points each. Those who are unemployed (mean=7.70, 95% CI: 7.18, 8.21) reported lower optimism than those who are employed by an outside organization (mean=8.14, 95% CI: 7.80, 8.47) or self-employed (mean=8.22, 95% CI: 7.93, 8.51). Moreover, optimism increased monotonically with years of education (e.g., mean_16+years_=8.23 [95% CI:7.92, 8.54] vs. mean _up to 8 years_=8.04 [95% CI: 7.68, 8.40]).

Participants who attend religious services frequently also reported higher mean levels of optimism. For instance, those who attended religious services more than once/week reported a mean optimism score of 8.54 (95% CI: 8.29, 8.79), which is 0.70 points higher than among those who never attended religious services (mean=7.84, 95% CI: 7.52, 8.17). Lastly, mean optimism is only slightly higher among the native-born (mean=8.11, 95% CI: 7.78, 8.43) than among immigrants (mean=8.08, 95% CI: 7.75, 8.41) by 0.03 points.

The calculated heterogeneity estimate τ (i.e. the estimated standard deviation of the means across countries) indicates that greater variation in mean optimism across countries is present within specific demographic categories, including among younger people (τ>0.90 in all age groups <40 years), those endorsing other gender identities (τ=1.61), those with domestic partners (τ=0.92), and people who are single/never married (τ=1.02) (Table 3). The calculated I^2^ is mostly above 95 across demographic categories, suggesting that variability in mean optimism across countries might be mainly due to true heterogeneity rather than sampling variability. The estimated prediction intervals are also wide for all sociodemographic categories, further providing evidence of heterogeneity across countries.

### Sociodemographic Variation in Optimism Levels: Country-Specific Estimates

In country-specific analyses, almost all demographic factors (except immigration status) are associated with optimism (global p-values <.05) in more than half of the countries, but the pattern varied widely across countries (Supplementary Table S1B to S22B; Supplementary Figures S1 to S34). For instance, optimism increased with age in almost all countries in North America, South America, and Europe (except Poland), whereas different patterns of associations were evident among African and Asian countries (e.g., optimism decreased with age in India but increased with age in Japan; optimism was not patterned by age in Kenya but decreased with age in Tanzania). In most countries, mean optimism is slightly higher in females vs. males; however, compared to both females and males, individuals identifying as other gender reported substantially lower optimism in many countries, even in countries with generally high social acceptance of gender minority populations (e.g., Sweden, United Kingdom, Australia, Germany)^51^; however, this estimate should be interpreted with caution as the imprecision with estimating these means are substantial, because the within-country sample size tended to be a very small proportion of the sample (≤1% in all countries). Married respondents also reported higher mean optimism than those who are divorced or single/never married and this pattern was evident in many countries, including in countries with low national divorce rates (e.g., India), high national divorce rates (e.g., Spain), and with prominent singlehood culture (e.g., Sweden)^52^. Moreover, participants with higher socioeconomic status also reported higher optimism levels in most countries. Specifically, mean optimism was higher among people who are employed vs. unemployed, even in countries with a good social welfare system (e.g., Sweden, Germany, United Kingdom). Similarly, higher optimism levels are correlated with more years of education in most countries. However, we observed substantial heterogeneity in the correlation between unemployment and optimism across countries (Figure S23). Overall, middle-income countries displayed relatively high mean optimism among unemployed people (e.g., mean=9.17 in Brazil, mean=9.05 in Mexico), whereas the unemployed in high-income countries reported low mean levels of optimism (e.g., mean=5.87 in Australia, mean=5.57 in Japan). In addition, individuals who reported attending religious services frequently tended to have higher optimism than those who never attended services, and this association was evident even in some of the most secular countries/territories^53^ (e.g., Japan, Hong Kong, Spain, Sweden).

Optimism differs by immigration status only in a handful of countries. Specifically, immigrants reported higher optimism in Australia, Spain, Sweden, United Kingdom, and United States, whereas the native-born had higher optimism in Brazil. Additional findings for optimism by religious affiliation and race/ethnicity (response categories for these two demographic factors varied across countries) are also presented with the country-specific analyses and reported in the supplement (Supplementary Table S1B to S22B).

The population weighted meta-analysis that pooled country-specific estimates considering population sizes in each country yielded largely similar results as the random effects meta-analysis, except that the mean optimism score in the oldest age groups were lower than that in the random effect meta-analyses (Supplementary Table S23). This is likely due to the heavy weight in the population weighted meta-analysis given to India where older people reported a low mean score of optimism. The complete-case analyses without imputation for missing data (Supplementary Tables S24 and S25) also yielded similar results as the primary analyses.

## DISCUSSION

Optimism is a psychological resource that has been shown to enhance health and well-being in some cultures^9^. However, whether this resource is similarly available to diverse populations is less well studied, and even studies considering the distribution of optimism across sociodemographic groups, have mainly been conducted in Western industrialized countries. The present study expands the literature by examining the mean levels of optimism across countries and also considering optimism’s distribution across major sociodemographic groups, with the samples weighted to be nationally representative of the adult population in each country.

Considering population levels of optimism across countries, this study finds both converging and diverging evidence from past research. Specifically, the mean scores of optimism are on the higher end in all countries in this sample (most countries have a mean score above 7 on a scale from 0-10), which is similar to prior findings^9,15^. This indicates that high levels of optimism may transcend geographic and cultural boundaries and optimism can potentially be achieved across diverse societal contexts. This study also finds that the rank ordering of optimism for many countries are similar to some earlier studies using data collected prior to 2010^9,15^ – for instance, Japan and Hong Kong have been consistently among the lower-ranking countries/regions, whereas Brazil, Mexico, and Nigeria have been consistently ranked high.

However, it is striking that several Western wealthy countries (e.g., United States, Sweden, United Kingdom, Australia) that showed high levels of optimism in earlier studies^9,15^ have the lowest rankings in the current sample (with data collected primarily in 2023). These somewhat contrary findings may be attributed to methodological differences as a number of previous studies often used student samples or assessed other aspects of positive psychological functioning (e.g., future life evaluation) as a proxy for optimism, whereas the current study relied upon samples of the adult population weighted to be nationally representative within each country and an item from a validated measure^33^ specifically designed for assessing optimism. Thus, it is possible that the findings may not be comparable between studies. Alternatively, if we assume the findings reflect true differences in optimism, such findings may indicate a declining trend of optimism in these wealthy countries over recent years, which may be related to the increased mental distress among some groups (e.g., younger people) in these societies over the past decade that was further exacerbated during the COVID-19 pandemic^54–57^. In addition, evidence from earlier studies indicated that optimism may be shaped by cultural values^9^ – e.g., optimism scores were often higher in countries that emphasize individualism and egalitarianism (e.g., Sweden, United States, United Kingdom, Australia). However, as these more individualistic countries show up among the lowest ranked countries in the present study, we need to consider whether factors beyond cultural values also matter. Other societal trends (e.g., technology development, economic prospects, community participation) that also differ cross-nationally may be playing a significant role in shaping optimism and well-being over recent years^56,58^. It is, however, also important to acknowledge that the observed differences in means of optimism between countries may be attributed to methodological complexities in cross-cultural survey research (e.g., translation of survey items, different response styles across cultures), thus the findings need to be interpreted with caution.

To our knowledge, this is the first cross-national study that calculated Gini coefficients to evaluate population inequality in optimism. In this sample countries that are considered collective tended to have more equal distribution in optimism, whereas countries that displayed the highest inequality in optimism are mostly high-income individualistic countries. Based on the World Bank’s estimates of income inequality by country^59^, population inequality in optimism and inequality in income do not seem to be strongly correlated. Overall, the findings may suggest that cultural values likely play a more important role than material resources in shaping overall equality in the distribution of optimism within societies. Future studies may consider using more nuanced measures of inequalities^60^ (e.g., absolute vs. relative measures of inequalities) to advance our understanding of inequality in optimism across populations, time, and settings.

The country specific analyses in this study add novel evidence on the age patterning of optimism within societies. Past studies suggested cross-national differences in the patterning of optimism by age in a limited number of countries– for instance, optimism increased with age in the United States, whereas the trend was reversed in Hong Kong^13^. The present study expands such evidence to a range of diverse countries, suggesting that optimism increases with age in most North American, South America, and European countries, whereas the pattern varied widely across Asian and African countries. It is striking that in this sample optimism among young people is considerably higher in economically underdeveloped countries than some of the richest countries, which is similar to findings from a recent survey by UNICEF^61^. Prior researchers hypothesize that this may be due to the dampened economic prospects and increased fear among young people in wealthy nations that they may become the first generation that does not do better than their parents economically^61,62^. Indeed, some wealthy nations have recently announced a mental health crisis among young people^60^. It is imperative to understand how to cultivate strengths (such as optimism) and reduce structural and social barriers for enhancing well-being among young people in these societies.

This study finds strong evidence of disparities in optimism by individuals’ socioeconomic status–-people who are employed (vs. unemployed) and people with more years of education reported greater optimism in most countries. These results are consistent with prior studies that mainly used data from Western countries^17,63,64^, but extend such evidence to a range of economically and culturally diverse countries. These findings may be understood with the Reserve Capacity Framework^65^, which posits that individuals with greater socioeconomic resources are less likely to encounter life stressors (e.g., job insecurity, housing instability, limited access to educational activities) and have greater capacity for coping with stressful encounters (e.g., greater access to mental health services, ability to afford childcare, flexibility to take time off work), leading to a more optimistic outlook for future outcomes even in difficult situations. In addition to the overall pattern between higher SES and higher optimism, this study also reveals important variations between countries. For instance, unemployment appeared to affect optimism more severely in individualistic countries than in collective countries.

Individualistic countries prioritize personal achievement and independence, and it is possible that people are more likely to integrate SES into their social identity and self-worth^66^. Thus, economic hardship and career challenges can lead to social stigma, a sense of personal failure, and loneliness; the high-living costs and competitive job markets in these societies may further intensify these challenges, which altogether negatively impacts individuals’ sense of optimism.

This study also expands evidence on greater religious participation and higher optimism to a wide range of countries. In this study participants who attend religious services frequently reported higher mean optimism levels, even in some of the most secular countries (e.g., Sweden, Spain, Japan). Such trends may be attributed in part to the practice of many faith traditions, especially the “salvation religions” (e.g., Christianity, Islam, Jainism, and Buddhism), which emphasizes promoting a sense of hope or optimism as an inherent component of their teaching^67^. In addition to strengthening faith, attending religious services also provides greater opportunities for social integration and connecting individuals to a larger community with shared beliefs and values who can also provide support during difficult times. Together, these practices and experiences can help further enhance optimism, belonging, and well-being^68^.

This study has some limitations. First, although optimism was assessed with an item from a commonly-used measure of optimism (LOT-R) that has been validated in multiple countries^69–71^, the GFS cognitive interviews found that some respondents had difficulty in understanding the optimism question/translation^28^, suggesting measurement error is possible. Moreover, the measure in this study does not distinguish different forms of optimism (e.g., whether the optimistic outlook for the future is grounded in one’s material resources or one’s personal agency in attaining positive future outcomes)^4^. Future studies that distinguish different forms of optimism will provide nuanced understanding of population distribution of optimism and how optimism may shape population well-being. Second, the country ordered means of optimism should be interpreted with caution, due to methodological challenges in cross-cultural survey research (e.g., survey question translation and interpretation, different response styles across cultures) and potential seasonal variation, as assessments were made in different countries during different times of the year. Because optimism was measured with a single item, we were not able to formally assess measurement invariance in optimism across countries. Third, there are important contextual factors that may shape optimism such as neighborhood conditions and within-country geographic variations that are not considered in this study due to lack of data.

Fourth, due to space constraints this study did not examine societal-level correlates of optimism, and the findings of this study may not be applicable to countries that are not included in the present study. Future studies that systematically examine societal-level conditions that shape optimism will deepen our understanding of whether, how, and why population levels of optimism vary across countries.

Having an optimistic mindset can be a desirable psychological attribute that may act as an asset for enhancing health and well-being in some cultures^9^. In this sample, population mean optimism varied across countries, and notably, some of the wealthiest countries had the lowest mean optimism levels, which warrants further investigation on societal determinants of optimism and their implications for population well-being. This study also provides novel evidence that optimism is not equally distributed across sociodemographic groups within individual countries. Some programs and interventions for enhancing optimism have been developed^72,73^. Preliminary evidence from randomized controlled trials suggests that such programs, when implemented appropriately, can be effective in promoting optimism and well-being outcomes^74^. It will be important to investigate culturally appropriate approaches to adapting such programs and to expanding access to interventions especially for vulnerable social groups. It is also important to gain greater insight into structural and social conditions that shape optimism and ways to enhance conditions that seem to promote higher population levels of optimism. Such activities will contribute to fostering population well-being and health equity, and to increasing resilience and flourishing across communities and societies.

## Acknowledgements

The Global Flourishing Study was supported by funding from the John Templeton Foundation (grant #61665), Templeton Religion Trust (#1308), Templeton World Charity Foundation (#0605), Well-Being for Planet Earth Foundation, Fetzer Institute (#4354), Well Being Trust, Paul L. Foster Family Foundation, and the David and Carol Myers Foundation. The funding source had no impact on the study design; on the collection, analysis and interpretation of data; on the writing of the report; or on the decision to submit the article for publication.

## Author Contributions

B.R. J., and T.J.V. developed the study concept. Y.C., L.D.K., E.S.K., H.K., K.S., R.N.P., R.W., B.R. J., and T.J.V. contributed to the study design. Y.C. and R.N.P. had full access to the data, conducted data analyses, and take responsibility for the integrity of the data and accuracy of the data analysis. Y.C. drafted the manuscript. L.D.K., E.S.K., H.K., K.S., R.N.P., R.W., B.R. J., and T.J.V. provided critical revisions, and approved the final submitted version of the manuscript.

## Ethics approval and consent to participate

Ethical approval was granted by the institutional review boards at Baylor University (IRB Reference #: 6431841317) and Gallup (IRB Reference #: 2021-11-02), and all participants provided informed consent.

## Data Availability

Data for Wave 1 of the Global Flourishing Study is available through the Center for Open Science upon submission of a pre-registration (https://doi.org/10.17605/OSF.IO/3JTZ8), and will be openly available without pre-registration beginning February 2025. Please see https://www.cos.io/gfs-access-data for more information about data access.

## Code Availability

All analyses were pre-registered with COS prior to data access (https://doi.org/10.17605/OSF.IO/WDSX2). All code to reproduce analyses are openly available in the online OSF repository (https://doi.org/10.17605/osf.io/vbype).

## Completing Interests

Tyler VanderWeele reports consulting fees from Gloo Inc., along with shared revenue received by Harvard University in its license agreement with Gloo according to the University IP policy. Other authors have no conflicts of interest to declare.

## Notes

### Author Declarations

Ethical approval was granted by the institutional review boards at Baylor University (IRB Reference #: 6431841317) and Gallup (IRB Reference #: 2021-11-02), and all participants provided informed consent. Data for Wave 1 of the Global Flourishing Study is available through the Center for Open Science upon submission of a pre-registration (https://doi.org/10.17605/OSF.IO/3JTZ8).

